## Supplementary figures and images for "Whole Genome Sequencing identifies novel structural variant in a large Indian family affected with X - linked agammaglobulinemia"

### Supplementary Figure 1

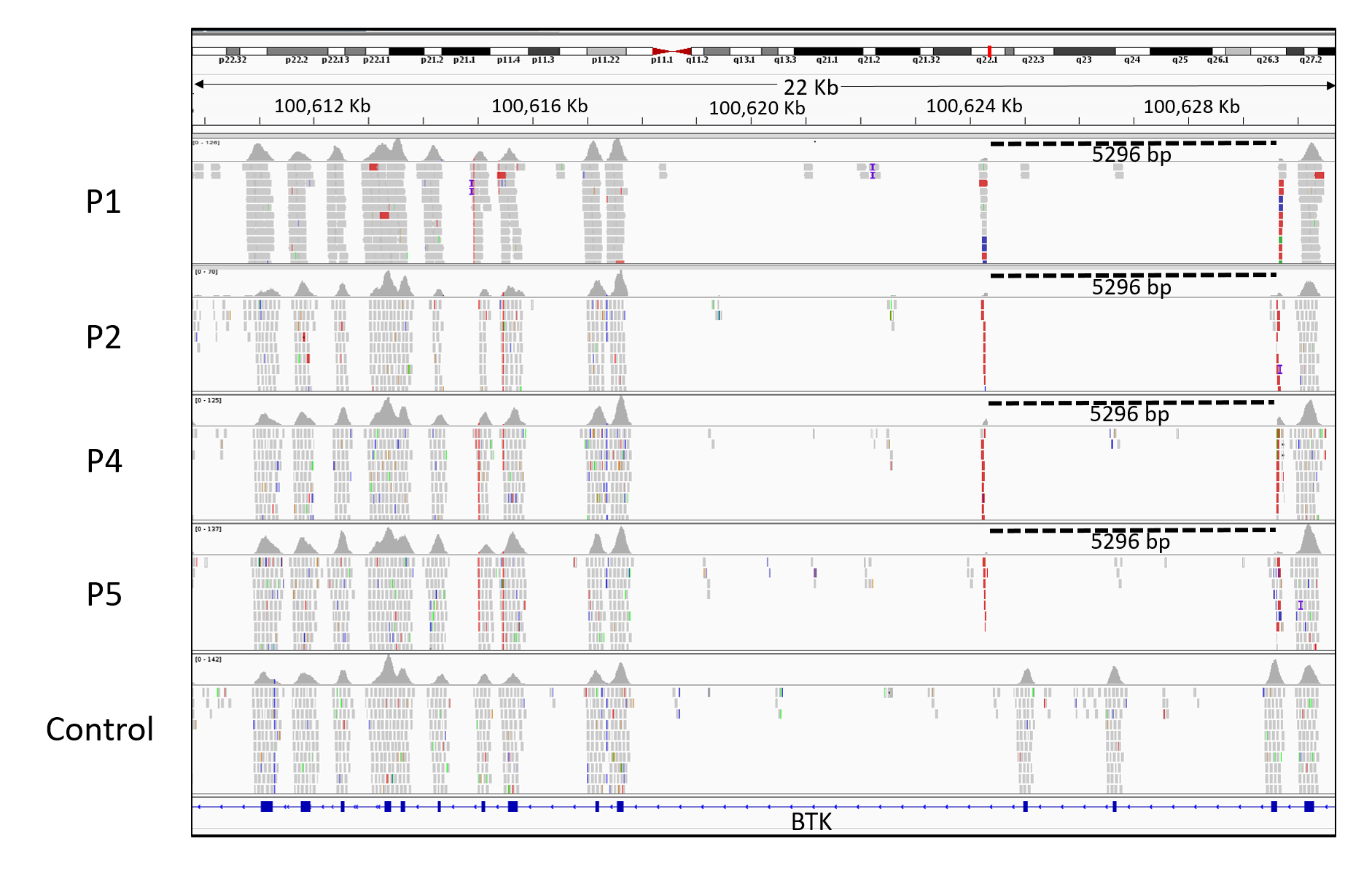

### Supplementary Figure 2

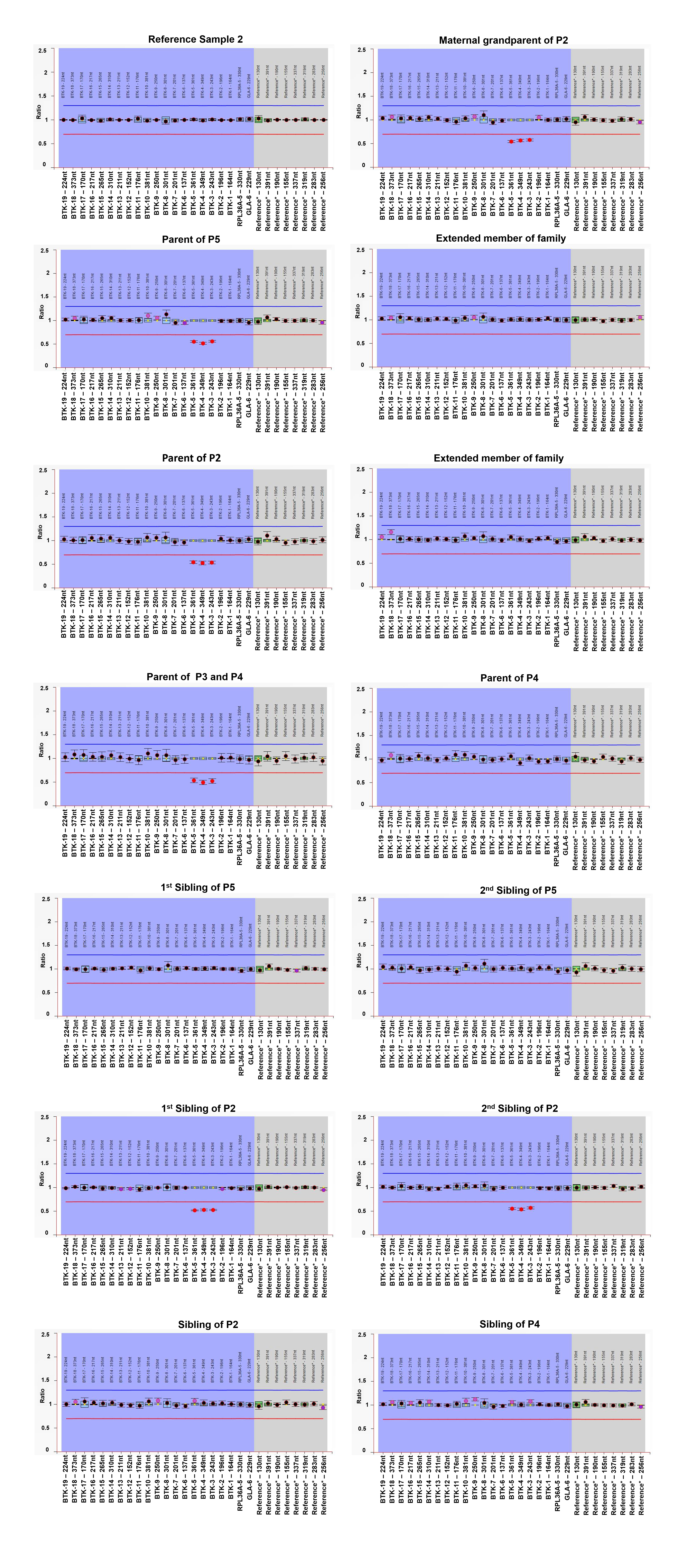

### Supplementary Figure 3

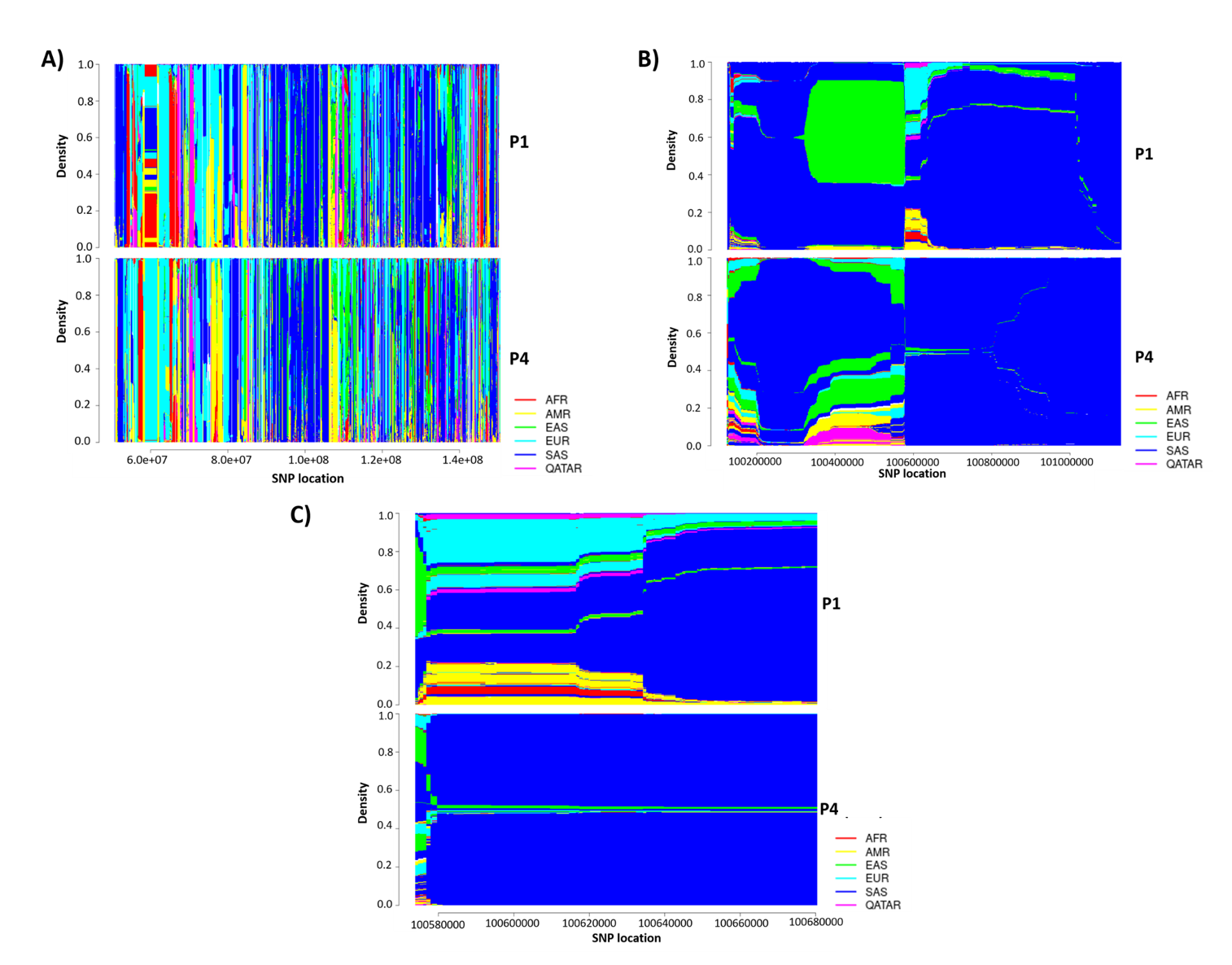
