## Supplementary Table 1 for "Whole Genome Sequencing identifies novel structural variant in a large Indian family affected with X - linked agammaglobulinemia"

| Data | P1 | P2 | P4 | P5 |
|---|---|---|---|---|
| **Total number of reads (x2)** | 39111154 | 41867264 | 37556352 | 41367110 |
| **Trimmed reads (x2)** | 22325405 | 31637375 | 18905832 | 36276744 |
| **Reads mapped (x2)** | 22320244 | 31616422 | 18851742 | 36276174 |
| **Percentage mapped** | 99.08% | 99.30% | 99.71% | 100% |
| **Coverage** | 74.4 | 105.4 | 62.8 | 120.9 |
| **Total variants** | 359,707 | 647,936 | 328,688 | 534,222 |
| **Protein altering variants** | 12,219 | 15,304 | 11,809 | 12,966 |
| **Rare variants (MAF < 5%)** | 1,552 | 4,185 | 2,193 | 1,602 |
| **PID genetic variants (454 gene)** | 9 | 9 | 17 | 16 |
| **Phenotype associated variant** | 0 | 0 | 0 | 0 |
| **Machine Used** | HiSeq2500 | HiSeq2500 | HiSeq2500 | NovaSeq6000 |
| **Library Used** | TruSeq DNA Exome | TruSeq DNA Exome | TruSeq DNA Exome | TruSeq DNA Exome |

**Supplementary Table 1:** Whole exome sequencing data summary for each patient.
