## Supplementary Table 2 for "Whole Genome Sequencing identifies novel structural variant in a large Indian family affected with X - linked agammaglobulinemia"

| Data | P1 | P4 |
|---|---|---|
| **Total number of reads** | 506,703,008 | 2,101,160,202 |
| **Trimmed reads** | 420,926,397 | 1,718,944,763 |
| **Reads mapped** | 419,160,298 | 1,710,162,053 |
| **Percentage mapped** | 99.58% | 99.49% |
| **Coverage** | 21 X | 86 X |
| **Total variants** | 4,905,687 | 4,904,615 |
| **Merged variants** | 6,517,075 | |
| **Rare variants (MAF < 5%)** | 890,387 | |
| **Common homozygous Variants** | 27,236 | |
| **In-silico prediction (CADD >15)** | 4 | |
| **Phenotype associated variant** | 0 | |
| **Machine Used** | NovaSeq6000 | NovaSeq6000 |
| **Library Used** | TruSeq PCR free | TruSeq PCR free |

**Supplementary Table 2:** Whole genome sequencing data summary for P1 and P4.
