## Supplementary Table 3 for "Whole Genome Sequencing identifies novel structural variant in a large Indian family affected with X - linked agammaglobulinemia"

| | | Reference sample 1 | Reference sample 2 | Maternal Grandparent of P2 | Parent of P5 | Sibling of parent of P1 | Parent of P2 | Sibling of parent of P1 | Parent of P3 and P4 | Parent of P3 and P4 | P5 | Sibling of P5 | Sibling of P5 | P1 | Sibling of P2 | Sibling of P2 | P2 | Sibling of P2 | Sibling of P3 and P4 | P4 |
|---|---|---|---|---|---|---|---|---|---|---|---|---|---|---|---|---|---|---|---|---|
| | BTK-19 - 224nt | 1 | 1 | 1.04 | 1.02 | 1.03 | 1.03 | 1.06 | 1.03 | 0.98 | 1.06 | 1.01 | 1.05 | 1.08 | 0.99 | 1.02 | 1.12 | 1 | 1.02 | 1.04 |
| | BTK-18 - 373nt | 1 | 1 | 1.07 | 1.05 | 1.05 | 1.01 | 1.16 | 1.08 | 1.08 | 1.05 | 0.99 | 1.03 | 1.04 | 1.02 | 1 | 0.98 | 1.06 | 1.06 | 0.97 |
| | BTK-17 - 170nt | 0.97 | 1.03 | 1.03 | 1.02 | 1.06 | 1.01 | 1.02 | 1.07 | 1.01 | 1.04 | 1 | 1.01 | 1 | 1 | 1.03 | 1.04 | 1.06 | 1.03 | 0.99 |
| | BTK-16 - 217nt | 1 | 1 | 1.04 | 1.01 | 1.03 | 1.06 | 1.02 | 1.03 | 1.01 | 1.06 | 1.01 | 1.04 | 1.11 | 1.01 | 1 | 1.11 | 1.04 | 1.05 | 1.06 |
| | BTK-15 - 265nt | 1.02 | 0.98 | 1.03 | 1.05 | 1.01 | 1.04 | 1 | 1.03 | 1.07 | 1.02 | 0.98 | 0.97 | 0.95 | 0.98 | 1.02 | 1.01 | 1.02 | 1.05 | 0.98 |
| | BTK-14 - 310nt | 0.99 | 1.01 | 1.06 | 1.05 | 1 | 1.06 | 0.99 | 1.07 | 1.02 | 1.09 | 1.01 | 0.99 | 1.04 | 0.99 | 0.97 | 1.03 | 1.03 | 1.07 | 0.96 |
| | BTK-13 - 211nt | 1 | 1 | 1.03 | 1 | 1.02 | 1 | 1.02 | 1.03 | 0.98 | 1 | 1.02 | 1.01 | 1 | 0.97 | 0.98 | 1.06 | 0.99 | 1 | 1 |
| | BTK-12 - 152nt | 1 | 1 | 1 | 1.01 | 1.02 | 0.98 | 1.01 | 0.99 | 1.02 | 1.02 | 1.03 | 1.01 | 1 | 0.97 | 1.02 | 1.07 | 0.98 | 0.99 | 1.05 |
| | BTK-11 - 176nt | 0.97 | 1.03 | 0.96 | 1.01 | 0.99 | 0.98 | 0.99 | 0.97 | 1.09 | 0.94 | 0.97 | 0.94 | 1.01 | 0.96 | 1.03 | 0.94 | 0.97 | 1.03 | 0.93 |
| | BTK-10 - 381nt | 1.02 | 0.98 | 1.03 | 1.1 | 1.04 | 1.07 | 1.07 | 1.11 | 1.09 | 1.05 | 1.01 | 1.07 | 1.07 | 0.99 | 1.05 | 1.02 | 1.06 | 1.09 | 1 |
| | BTK-9 - 250nt | 1 | 1 | 1.07 | 1.06 | 1.06 | 1.06 | 1.03 | 1.07 | 1.05 | 1.11 | 1 | 1.02 | 1.08 | 1 | 1.03 | 1.04 | 1.08 | 1.12 | 0.97 |
| | BTK-8 - 301nt | 1.03 | 0.97 | 1.1 | 1.13 | 1.06 | 1.07 | 1.07 | 1.07 | 0.99 | 1.12 | 1.08 | 1.11 | 1.08 | 1.01 | 1.04 | 1.11 | 1.03 | 1.05 | 1.1 |
| | BTK-7 - 201nt | 1.01 | 0.99 | 0.95 | 0.95 | 1.01 | 0.97 | 1.01 | 0.97 | 0.98 | 0.96 | 1.01 | 1 | 1.01 | 1 | 0.97 | 1.01 | 0.97 | 0.97 | 0.98 |
| | BTK-6 - 137nt | 1 | 1 | 0.99 | 0.95 | 0.99 | 0.96 | 0.99 | 0.98 | 1 | 0.96 | 1.02 | 1.02 | 1.01 | 1 | 1 | 1.05 | 0.96 | 0.98 | 1.01 |
| | BTK-5 - 361nt | 0.99 | 1.01 | 0.54 | 0.55 | 1.01 | 0.54 | 1.05 | 0.54 | 1.06 | 0 | 1 | 1.01 | 0 | 0.52 | 0.56 | 0 | 1.08 | 1.06 | 0 |
| | BTK-4 - 349nt | 1.01 | 0.99 | 0.57 | 0.52 | 1.02 | 0.53 | 1.01 | 0.49 | 0.92 | 0 | 1.01 | 1.03 | 0 | 0.53 | 0.54 | 0 | 1.03 | 1.01 | 0 |
| | BTK-3 - 243nt | 0.99 | 1.01 | 0.58 | 0.56 | 1 | 0.54 | 0.97 | 0.52 | 1.02 | 0 | 1.02 | 0.98 | 0 | 0.53 | 0.57 | 0 | 1.02 | 1.08 | 0 |
| | BTK-2 - 196nt | 1 | 1 | 1.05 | 0.99 | 1.01 | 1.04 | 1.02 | 1.01 | 0.95 | 1.05 | 1 | 1.03 | 1.1 | 0.97 | 0.99 | 1.11 | 1.02 | 1.02 | 1.04 |
| | BTK-1 - 164nt | 1 | 1 | 1 | 0.99 | 1.03 | 1.01 | 1.03 | 1.02 | 0.95 | 1.03 | 1 | 1.03 | 1.05 | 0.99 | 0.98 | 1.08 | 1.01 | 1 | 1.04 |
| | RPL36A-5 - 330nt | 0.98 | 1.02 | 1.01 | 1 | 1 | 1 | 0.95 | 0.99 | 1 | 0.92 | 0.96 | 0.95 | 1.07 | 0.97 | 0.98 | 0.93 | 1 | 1.02 | 0.95 |
| **Test probes** | GLA-6 - 229nt | 0.98 | 1.02 | 1.02 | 0.96 | 1 | 1.03 | 0.97 | 0.97 | 0.97 | 0.96 | 1 | 0.97 | 1.01 | 0.98 | 0.99 | 1.01 | 0.99 | 0.98 | 0.99 |
| | Reference - 130nt | 0.97 | 1.03 | 0.95 | 0.98 | 1 | 0.98 | 0.99 | 0.94 | 1.05 | 0.95 | 0.98 | 0.93 | 1.05 | 0.97 | 1 | 0.95 | 0.96 | 1.01 | 0.98 |
| | Reference - 391nt | 1.01 | 0.99 | 1.06 | 1.07 | 1.01 | 1.11 | 1.06 | 1.04 | 0.98 | 1.09 | 1.06 | 1.06 | 1.03 | 1.02 | 0.99 | 1.05 | 1.02 | 1.07 | 1.04 |
| | Reference - 190nt | 1 | 1 | 1.01 | 1.03 | 1.02 | 1.03 | 1.03 | 0.95 | 0.95 | 1.03 | 1 | 1.02 | 1.05 | 1 | 1.01 | 1.02 | 0.99 | 1 | 1.02 |
| | Reference - 155nt | 1 | 1 | 0.99 | 1 | 0.97 | 0.95 | 0.98 | 1.05 | 1.05 | 1 | 0.98 | 0.97 | 0.98 | 1.01 | 0.99 | 1 | 1.04 | 1 | 0.97 |
| | Reference - 337nt | 1 | 1 | 0.98 | 1 | 1 | 0.98 | 1.01 | 0.99 | 1.01 | 0.98 | 0.97 | 0.96 | 0.93 | 0.99 | 1.03 | 0.97 | 1 | 0.99 | 0.99 |
| | Reference - 319nt | 0.99 | 1.01 | 1.02 | 1.01 | 0.99 | 1 | 0.98 | 1.02 | 0.95 | 1.03 | 1.02 | 1.03 | 1.03 | 1.01 | 0.97 | 1.05 | 1 | 0.99 | 1.01 |
| | Reference - 283nt | 1 | 1 | 1.02 | 1 | 1 | 0.99 | 1 | 1.02 | 1.02 | 1 | 1 | 1 | 0.98 | 1 | 0.98 | 1 | 1.01 | 1.02 | 0.98 |
| **Reference probes** | Reference - 256nt | 1 | 1 | 0.95 | 0.96 | 1.05 | 1.01 | 0.99 | 0.95 | 0.99 | 0.94 | 0.99 | 1 | 0.98 | 0.95 | 1.02 | 0.97 | 0.93 | 0.96 | 1 |
